# Lowered oxygen saturation and increased body temperature in acute COVID-19 largely predict chronic fatigue syndrome and affective symptoms due to LONG COVID: a precision nomothetic approach

**DOI:** 10.1101/2022.04.10.22273660

**Authors:** Dhurgham Shihab Al-Hadrawi, Haneen Tahseen Al-Rubaye, Abbas F. Almulla, Hussein Kadhem Al-Hakeim, Michael Maes

## Abstract

**Background:** Long coronavirus disease 2019 (LC) is a chronic sequel of acute COVID-19. The exact pathophysiology of the affective, chronic fatigue and physiosomatic symptoms (labeled as “physio-affective phenome”) of LC has remained elusive.

**Objective:** The current study aims to delineate the effects of oxygen saturation (SpO2) and body temperature during the acute phase on the physio-affective phenome of LC.

**Method:** We recruited 120 LC patients and 36 controls. For all participants, we assessed the lowest SpO2 and peak body temperature during acute COVID-19, and the Hamilton Depression and Anxiety Rating Scale (HAMD/HAMA) and Fibro Fatigue (FF) scales 3 to 4 months later.

**Results:** Lowered SpO2 and increased body temperature during the acute phase and female sex predict 60.7% of the variance in the physio-affective phenome of LC. Using unsupervised learning techniques we were able to delineate a new endophenotype class, which comprises around 26.7% of the LC patients and is characterized by very low SpO2 and very high body temperature, and depression, anxiety, chronic fatigue, and autonomic and gastro-intestinal symptoms scores. Single latent vectors could be extracted from both biomarkers, depression, anxiety and FF symptoms or from both biomarkers, insomnia, chronic fatigue, gastro-intestinal and autonomic symptoms.

**Conclusion:** The newly constructed endophenotype class and pathway phenotypes indicate that the physio-affective phenome of LC is at least in part the consequence of the pathophysiology of acute COVID-19, namely the combined effects of lowered SpO2, increased body temperature and the associated immune-inflammatory processes and lung lesions.

## Introduction

Long coronavirus disease 2019 or post-corona virus disease 2019 (post-COVID-19 or Long COVID) is a sequel of prior infection with severe acute respiratory syndrome coronavirus 2 (SARS-CoV-2) (Nalbandian, Sehgal et al. 2021, World Health Organization 2022). This syndrome is manifested as a cluster of symptoms mainly but not limited to fatigue, shortening of breath, persistent cough, chest pain, cognitive impairments, and affective symptoms (Renaud-Charest, Lui et al. 2021, Sandler, Wyller et al. 2021, Titze-de-Almeida, da Cunha et al. 2022). Similar consequences were also reported in previous epidemics, for example SARS-2003 and the Middle East respiratory syndrome (MERS-2012) (Lam, Wing et al. 2009, Moldofsky and Patcai 2011, Lee, Shin et al. 2019, Ahmed, Patel et al. 2020).

There is a growing concern that Long COVID is becoming a serious health issue (Phillips and Williams 2021). Six months after the acute infection, 33% of COVID-19 patients may experience serious neuropsychiatric symptoms, while 13% of them even received a first diagnosis months after the acute phase (Taquet, Geddes et al. 2021). Regardless of whether COVID-19 patients were symptomatic or asymptomatic during the acute phase of illness, 10-20% of them will experience Long COVID symptoms within weeks to months after recovery (Huang, Pinto et al. 2021, World Health Organization 2022). Other results show that 80% of the recovered COVID- 19 patients suffer from at least one of the Long COVID symptoms, including fatigue, memory impairment, anxiety and depression (Lopez-Leon, Wegman-Ostrosky et al. 2021, Badenoch, Rengasamy et al. 2022). Interestingly, the prevalence of Long COVID is not affected by hospitalization status, disease severity, or length of follow-up (Davido, Seang et al. 2020, Badenoch, Rengasamy et al. 2022).

Acute SARS-CoV-2 infection is characterized by an exaggerated immune-inflammatory response and infiltration of the inflammatory mediators including pro-inflammatory cytokines into the lung tissues (Mehta, McAuley et al. 2020, Pelaia, Tinello et al. 2020, Al-Jassas, Al-Hakeim et al. 2022). The consequent lung injuries, which may be identified by chest computerized tomography abnormalities (CCTAs), are accompanied by lowered oxygen saturation (SpO2) which may aggravate the inflammatory responses and may persist even after full recovery (Solomon, Heyman et al. 2021, Vijayakumar, Tonkin et al. 2021, Al-Jassas, Al-Hakeim et al. 2022). Increased body temperature in the acute phase of illness is one of the most common signs of infection and inflammation and this marker is widely used to detect febrile SARS-CoV-2 individuals (Lippi et al., 2021). The degree of increments in body temperature reflect the severity of inflammation and the peak body temperature during the acute phase is associated with an increased mortality risk (Tharakan, Nomoto et al. 2020).

The onset of Long COVID is attributed to precipitating factors associated with SARS-CoV-2 infection including abnormal immune responses, inflammatory damage, alterations in microbiome/virome in response to viral interactions, hypercoagulability, abnormal signaling of the brainstem and vagus nerve, and even physical adaptations to inactivity or psychological factors (Calabrese 2020, Deng, Zhou et al. 2021, Nalbandian, Sehgal et al. 2021, Proal and VanElzakker 2021). Furthermore, the onset of Long COVID fatigue was attributed to predisposing genetic and psychosocial vulnerabilities, and its socio-economic consequences, and perpetuating factors such as sleep disturbances, autonomic dysfunctions, and aberrations in endocrine functions (Theorell, Blomkvist et al. 1999, Papadopoulos and Cleare 2011, Jackson and Bruck 2012, Piraino, Vollmer-Conna et al. 2012, Cvejic, Li et al. 2019, Nelson, Bahl et al. 2019, Sandler, Wyller et al. 2021). Moreover, SARS-CoV-2 infected people may show long-term effects on brain structure and functions (Boldrini, Canoll et al. 2021), which may be due to neuroinflammation or the direct effect of hypoxia (Solomon 2021, Song, Zhang et al. 2021).

Nonetheless, no studies examined the effects of acute COVID-19 biomarkers, such as lowered SpO2 and increased body temperature, on the mental and chronic fatigue symptoms during Long COVID. Hence, the aim of this study is to delineate the effects of SpO2 and body temperature during the acute phase on chronic fatigue syndrome and affective symptoms in Long COVID. In the current study, we use the precision nomothetic approach (Maes 2022) to delineate new pathway phenotypes and endophenotype classes which combine those two infection biomarkers with Long COVID mental and chronic fatigue symptoms. Such data are needed to understand the pathophysiology of Long COVID and post-viral symptoms in general and may help to predict who will develop chronic fatigue syndrome and affective symptoms due to COVID-19 and viral infections in general.

## Participants and Methods

### Participants

In the present study, we used a case-control study design (to examine differences between controls and Long COVID subtypes) as well as a retrospective cohort study design (to examine the effects of acute phase biomarkers on Long COVID symptoms). During the last three months of 2021, we recruited 120 participants who suffered from at least 2 symptoms of Long COVID and who were previously diagnosed and treated for acute COVID-19 infection. During their acute phase, the Long COVID participants had been admitted to various hospitals and centers in Al-Najaf city for treatment of acute COVID-19, namely Al-Sader Medical City of Najaf, Al-Hakeem General Hospital, Al-Zahraa Teaching Hospital for Maternity and Pediatrics, Imam Sajjad Hospital, Hassan Halos Al-Hatmy Hospital for Transmitted Diseases, Middle Euphrates Center Cancer, Al-Najaf Center for Cardiac Surgery and Trans Catheter Therapy. All patients had been diagnosed as moderate to severe acute COVID-19 based on their clinical symptoms and the WHO criteria (World Health Organization 2022) and positive results of reverse transcription real-time polymerase chain reaction (rRT-PCR). Upon recovery all patients showed a negative rRT-PCR test. Three to four months after admission for acute COVID-19 they showed at least two symptoms that were present for at least two months including fatigue, memory or concentration disorders, shortness of breath or difficulty breathing, chest pain, persistant cough, trouble speaking, muscle aches, loss of smell or taste, affective symptoms or fever (World Health Organization 2022). Additionally, we recruited 36 controls from the same catchment area, who were either employees or family or friends of staff members. We also included controls who demonstrated distress or adjustment symptoms because of lockdowns and social isolation to account for their confounding effects that are also evident in Long COVID patients. As such, 1/3 of the controls show HAMD levels between 7 and 12. All controls showed a negative rRT-PCR test and no clinical signs of acute infection including dry cough, sore throat, shortness of breath, loss of appetite, flu-like symptoms, fever, night sweats, and chills. Patients and controls were excluded if they had a lifetime history of psychiatric disorders, including major depression, bipolar disorder, anxiety disorders, schizophrenia, and substance use disorders, except tobacco use disorder (TUD), neuroinflammatory or neurodegenerative disorders including multiple sclerosis, chronic fatigue syndrome (Morris and Maes 2013), Parkinson’s and Alzheimer’s disease, and stroke, and systemic (auto)immune diseases such as diabetes mellitus, COPD, rheumatoid arthritis and psoriasis, and liver and renal diseases. We also excluded pregnant and lactating women.

Before participating in the study, all controls and patients or their parents/legal guardians provided written signed consent. The approval of the study was obtained from the institutional ethics board of the University of Kufa (617/2020). The study was accomplished under Iraqi and foreign ethics and privacy rules according to the guidelines of the World Medical Association Declaration of Helsinki, The Belmont Report, CIOMS Guideline, and International Conference on Harmonization of Good Clinical Practice; our IRB adheres to the International Guideline for Human Research Safety (ICH-GCP).

### Clinical assessments

A well-trained paramedical professional recorded spO2 with an electronic oximeter provided by Shenzhen Jumper Medical Equipment Co. Ltd. and body temperature as assessed using a digital oral thermometer (sublingual until the beep). In the present study, we extracted both biomarkers from the patient records and used the lowest SpO2 and peak body temperature data that were measured during the acute phase of illness in the analyses. Based on those two assessments we computed a new index which reflects lowered SpO2 and higher temperature as z transformation of body temperature (z T) - z SpO2 (named the “TO2 index”). In all participants we registered the vaccinations they had received, namely AstraZeneca, Pfizer or Sinopharm. A semi-structured interview, conducted by a senior psychiatrist, assessed sociodemographic and clinical data in controls and Long COVID patients three to four months after recovery (mean ±SD: 14.68 ±5.31 weeks) from acute COVID-19. We assessed the following rating scales: a) depressive symptoms were examined utilizing the 21-item Hamilton Depression Rating Scale (HDRS) score (Hamilton 1960); b) anxiety symptoms were assessed using the Hamilton Anxiety Rating Scale (HAM-A) (Hamilton 1959); and c) and chronic fatigue and fibromyalgia symptoms using the Fibro-Fatigue (FF) 12-item scale (Zachrisson, Regland et al. 2002).

We computed two HAMD subdomain scores: a) pure depressive symptoms (pure HAMD) were calculated as the sum of depressed mood + feelings of guilt + suicidal ideation + loss of interest; and b) physiosomatic HAMD symptoms (Physiosom HAMD) was computed as: anxiety somatic + gastrointestinal + genitourinary + hypochondriasis. Two HAMA subdomain scores were computed: a) key anxiety symptoms (Key HAMA) as anxious mood + tension + fears + anxiety behavior at interview; and b) physiosomatic HAMA symptoms (Physiosom HAMA) as somatic sensory + cardiovascular + gastrointestinal (GIS)+ genitourinary + autonomic symptoms (respiratory symptoms were not included in the sum). We computed one pure physiosom FF subdomain score as muscle pain + muscle tension + fatigue + autonomous symptoms + gastrointestinal symptoms + headache + a flu-like malaise (thus excluding the cognitive and affective symptoms). Moreover, using all relevant HAMD, HAMA, and FF items (z transformed) we calculated z unit based composite scores reflecting autonomic symptoms, sleep disorders, fatigue, gastro-intestinal symptoms, and cognitive symptoms. We calculated the body mass index (BMI) based on the equation dividing body weight in kilograms by height in meter ^2^. We made the diagnosis of TUD using DSM-5 criteria.

### Data analysis

Differences in continuous variables between groups were checked using analysis of variance (ANOVA). Analysis of contingency tables (the χ2-test) was used to determine the association between nominal variables. Correlations between two variables were assessed using Pearson’s product moment correlation coefficients. We employed multivariate and univariate general linear model (GLM) analysis to delineate the associations between study groups (controls versus patients divided into those with low and high TO2 index scores) and rating scale scores while controlling for confounding variables including age, sex, smoking and education. Consequently, we computed the estimated marginal mean values (SE) and conducted protected (the omnibus test is significant) LSD tests to conduct pairwise comparisons among the group means. Multiple comparisons were subjected to false discovery rate (FDR) p-correction (Benjamini and Hochberg 1995). Moreover, we used multiple regression analysis to delineate significant predictors of the rating scale scores while allowing for the effects of confounders. An automated stepwise method was employed with an 0.05 p-value to entry and 0.06 to remove. We computed for each significant explanatory variable the standardized beta coefficients with t statistics and exact p-value, and for the model F statistics and total variance explained (R^2^). Moreover, we always checked changes in R^2^ and collinearity issues using the variance inflation factor and tolerance. The White and modified Breusch-Pagan tests for homoscedasticity were used to check heteroskedasticity and if needed we computed the parameter estimates with robust errors using univariate GLM analysis. The significance was determined at p=0.05, and two-tailed tests were applied. Power analysis showed that using an effect size of 0.23, p=0.05, power=0.8 and three groups with up to 5 covariates in an analysis of variance the sample size should be around 151 subjects. Therefore, we included 156 subjects, namely 36 controls and 120 Long COVID participants.

In accordance with the precision nomothetic approach (Maes 2022) we aimed to construct endophenotype classes of Long COVID patients (using cluster analysis), and new pathway phenotypes (using factor analysis) by combining biomarker and clinical data. Exploratory factor analysis (unweighted least squares extraction, 25 iterations for convergence) was performed, and the Kaiser-Meier-Olkin (KMO) sample adequacy measure was used to assess factorability (sufficient when >0.7). Moreover, when all loadings on the first factor were > 0.6 and the variance explained by the first factor was > 50.0%, and Cronbach alpha performed on the variables was > 0.7, the first PC was regarded as a valid latent construct underpinning the variables. Canonical correlation analysis was used to examine the relationships between two sets of variables, whereby symptoms three to four months after the acute phase were entered as dependent variables and the biomarkers as explanatory variables. We computed the variance explained by the canonical variables of both sets and the variance in the canonical dependent variables set explained by the independent canonical variable set. The canonical components are accepted when the explained variance of both sets is > 0.5 and when all canonical loadings are > 0.5. Two step cluster analysis was performed considering categorical and continuous variables. The cluster solution was considered adequate when the silhouette measure of cohesion and separation was > 0.5. IBM SPSS windows version 28 was used for all statistical analyses.

## Results

### Sociodemographic data

In order to divide the patient sample in two subgroups based on baseline SpO2 and body temperature data we performed two-step cluster analysis with being infected or not as categorical variable and body temperature and SpO2 as continuous variables. This cluster analysis showed three clusters with adequate cluster quality (silhouette measure of cohesion and separation of 0.62) comprising the healthy control sample (n=36), and patients with a low (group 1, n=88) versus very high (group 2, n=32) TO2 index. As such, patients with Long COVID were divided according to measurements during the acute infectious phase. **Table 1** shows the sociodemographic data of these three groups. Group 2 patients (high TO2 index) showed a significant increase in body temperature and decreased spO2 values as compared to group 1 patients (low TO2 index) and controls, while the low TO2 group showed lower SpO2 and higher temperature than controls. No significant differences in these groups were found in sex, TUD, residency, vaccination status, and BMI. The mean age was somewhat higher and education somewhat lower in the high TO2 group as compared with the low TO2 group and controls.

**Table 1.** Socio-demographic data, body temperature (BT) and oxygen saturation (SpO2) in control participants (CP) and Long COVID (LC) patients divided according to their TO2 index.

| Variables | CP (n=36) <sup>A</sup> | LC and lower TO<sub>2</sub> (n=88) <sup>B</sup> | LC and high TO<sub>2</sub> (n=32) <sup>C</sup> | F/KWT/X<sup>2</sup> | df | p |
| --- | --- | --- | --- | --- | --- | --- |
| Age (years) | 30.9 (8.3) <sup>C</sup> | 29.7 (7.3) <sup>C</sup> | 35.6 (9.6) <sup>A,B</sup> | 6.22 | 2/153 | 0.003 |
| Sex (M/F) | 30/6 | 59/29 | 26/6 | 4.67 | 2 | 0.096 |
| Marital state (Ma/S) | 14/22 | 48/40 | 23/9 | 7.43 | 2 | 0.024 |
| Smoking (Y/N) | 16/20 | 28/60 | 9/23 | 2.43 | 2 | 0.297 |
| Residency (U/R) | 29/7 | 72/16 | 29/3 | 1.57 | 2 | 0.456 |
| Vaccination (A/PF/S) | 11/14/11 | 37/34/17 | 6/17/9 | 6.86 | 4 | 0.145 |
| BMI (kg/m<sup>2</sup>) | 26.3 (3.6) | 26.3 (5.1) | 26.1 (5.4) | 0.03 | 2/148 | 0.975 |
| Education (years) | 15.8 (1.2) <sup>C</sup> | 15.8 (1.7) <sup>C</sup> | 14.9 (1.3) <sup>A,B</sup> | 3.61 | 2/153 | 0.029 |
| Maximal BT (°C) | 36.5 (0.1) <sup>B,C</sup> | 38.7 (0.5) <sup>A,C</sup> | 40.1 (0.7) <sup>A,B</sup> | KWT | - | <0.0001 |
| Lowest SpO<sub>2</sub> (%) | 96.58 (1.48) <sup>B,C</sup> | 91.50 (3.06) <sup>A,C</sup> | 85.84 (6.30) <sup>A,B</sup> | KWT | - | <0.0001 |
| TO<sub>2</sub> index (z scores) | -1.338 (0.179) <sup>B,C</sup> | 0.155 (0.398) <sup>A,C</sup> | 1.345 (0.659) <sup>A,B</sup> | KWT | - | <0.0001 |

Results are shown as mean (SD): F: results of analysis of variance; KWT: Kruskal Wallis test; X<sup>2</sup>: analysis of contingency tables.
M: Male, F: Female, Ma: Married, S: Single, U: Urban, R: Rural, BMI: Body Mass Index, TO<sub>2</sub> index: computed as z BT – z SpO<sub>2</sub>.
A: AstraZeneca, Pf: Pfizer, S: Sinopharm
<sup>A,B,C</sup>: Results of pairwise comparisons among group means

### Differences in psychiatric rating scales between study groups

The measurements of the total and subdomains scores of the rating scales are displayed in **Table 2**. All total scores, the pure and physiosom HAMD and HAMA and pure FF scores and severity of autonomic and gastro-intestinal symptoms were significantly different between the three study groups and increased from controls → low TO2 group → high TO2 group. Furthermore, there were significant differences in pure HAMA, sleep disorders, fatigue and cognitive impairments between Long COVID patients and controls with a trend toward higher values in the high TO2 group. The intergroup differences remained significant using an FDR of p=0.01. Consequently, we have extracted the first factor from the pure and physiosom HAMD and HAMA and pure FF scores (this first factor explained 66.99% of the variance; KMO=0.877, all loadings on the first factor > 0.724). This factor therefore underpins the different subdomains and was labeled the “physio-affective core” or “physio-affective phenome” of Long COVID. Table 2 shows that this score was significantly different between the three groups.

**Table 2.**
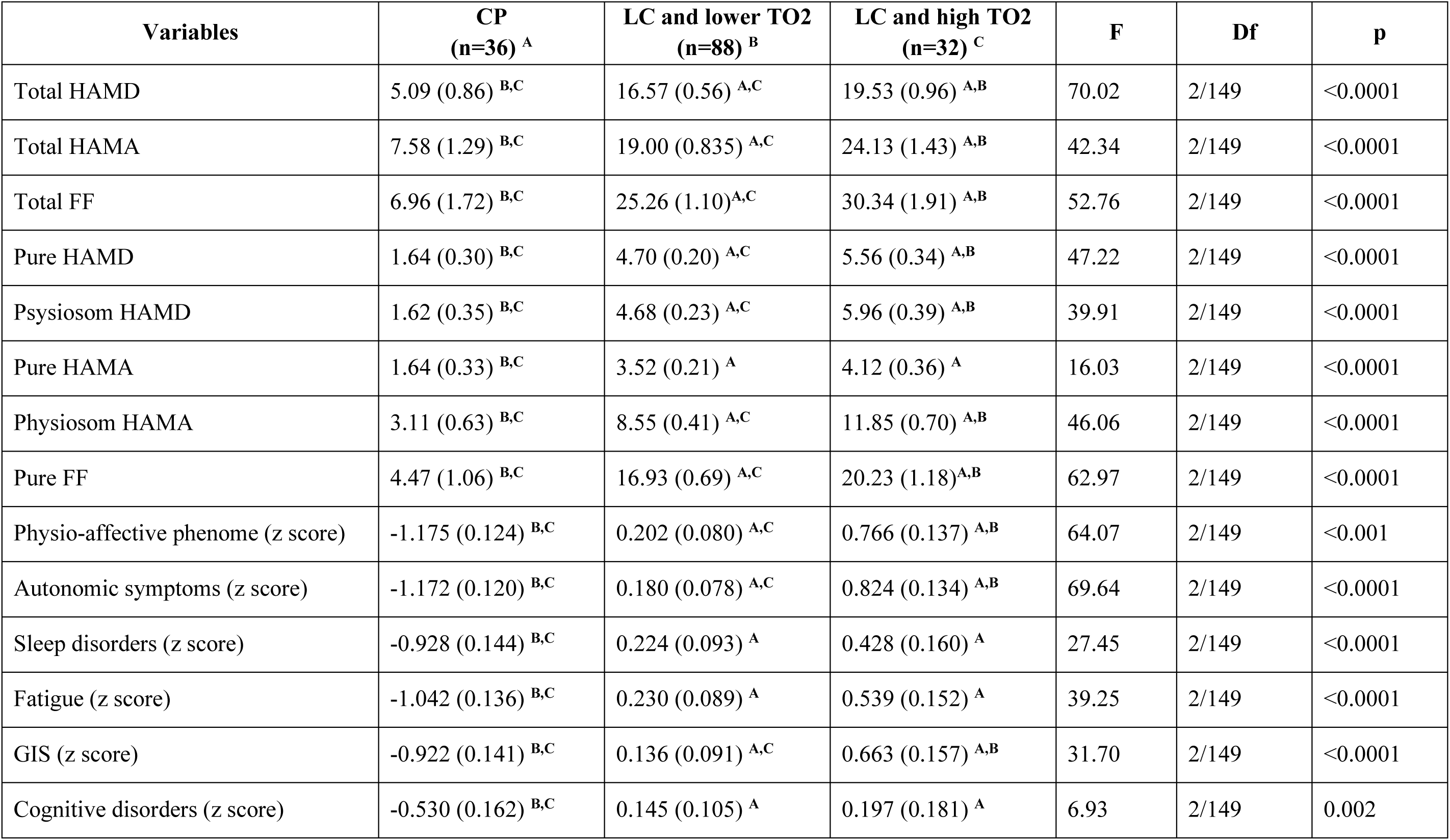
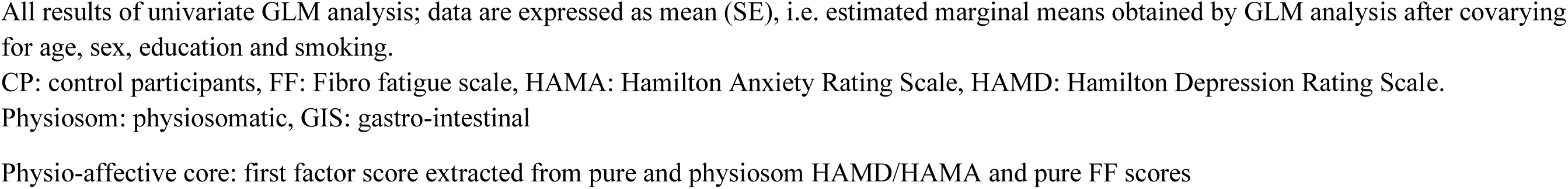
Clinical rating scales scores in control participants (CP) and Long COVID (LC) patients divided according to their TO2 index.

| **Variables** | **CP**<br>**(n=36) <sup>A</sup>** | **LC and lower TO2**<br>**(n=88) <sup>B</sup>** | **LC and high TO2**<br>**(n=32) <sup>C</sup>** | **F** | **Df** | **p** |
| --- | --- | --- | --- | --- | --- | --- |
| Total HAMD | 5.09 (0.86) <sup>B,C</sup> | 16.57 (0.56) <sup>A,C</sup> | 19.53 (0.96) <sup>A,B</sup> | 70.02 | 2/149 | <0.0001 |
| Total HAMA | 7.58 (1.29) <sup>B,C</sup> | 19.00 (0.835) <sup>A,C</sup> | 24.13 (1.43) <sup>A,B</sup> | 42.34 | 2/149 | <0.0001 |
| Total FF | 6.96 (1.72) <sup>B,C</sup> | 25.26 (1.10) <sup>A,C</sup> | 30.34 (1.91) <sup>A,B</sup> | 52.76 | 2/149 | <0.0001 |
| Pure HAMD | 1.64 (0.30) <sup>B,C</sup> | 4.70 (0.20) <sup>A,C</sup> | 5.56 (0.34) <sup>A,B</sup> | 47.22 | 2/149 | <0.0001 |
| Psysiosom HAMD | 1.62 (0.35) <sup>B,C</sup> | 4.68 (0.23) <sup>A,C</sup> | 5.96 (0.39) <sup>A,B</sup> | 39.91 | 2/149 | <0.0001 |
| Pure HAMA | 1.64 (0.33) <sup>B,C</sup> | 3.52 (0.21) <sup>A</sup> | 4.12 (0.36) <sup>A</sup> | 16.03 | 2/149 | <0.0001 |
| Physiosom HAMA | 3.11 (0.63) <sup>B,C</sup> | 8.55 (0.41) <sup>A,C</sup> | 11.85 (0.70) <sup>A,B</sup> | 46.06 | 2/149 | <0.0001 |
| Pure FF | 4.47 (1.06) <sup>B,C</sup> | 16.93 (0.69) <sup>A,C</sup> | 20.23 (1.18) <sup>A,B</sup> | 62.97 | 2/149 | <0.0001 |
| Physio-affective phenome (z score) | -1.175 (0.124) <sup>B,C</sup> | 0.202 (0.080) <sup>A,C</sup> | 0.766 (0.137) <sup>A,B</sup> | 64.07 | 2/149 | <0.001 |
| Autonomic symptoms (z score) | -1.172 (0.120) <sup>B,C</sup> | 0.180 (0.078) <sup>A,C</sup> | 0.824 (0.134) <sup>A,B</sup> | 69.64 | 2/149 | <0.0001 |
| Sleep disorders (z score) | -0.928 (0.144) <sup>B,C</sup> | 0.224 (0.093) <sup>A</sup> | 0.428 (0.160) <sup>A</sup> | 27.45 | 2/149 | <0.0001 |
| Fatigue (z score) | -1.042 (0.136) <sup>B,C</sup> | 0.230 (0.089) <sup>A</sup> | 0.539 (0.152) <sup>A</sup> | 39.25 | 2/149 | <0.0001 |
| GIS (z score) | -0.922 (0.141) <sup>B,C</sup> | 0.136 (0.091) <sup>A,C</sup> | 0.663 (0.157) <sup>A,B</sup> | 31.70 | 2/149 | <0.0001 |
| Cognitive disorders (z score) | -0.530 (0.162) <sup>B,C</sup> | 0.145 (0.105) <sup>A</sup> | 0.197 (0.181) <sup>A</sup> | 6.93 | 2/149 | 0.002 |

All results of univariate GLM analysis; data are expressed as mean (SE), i.e. estimated marginal means obtained by GLM analysis after covarying for age, sex, education and smoking.
CP: control participants, FF: Fibro fatigue scale, HAMA: Hamilton Anxiety Rating Scale, HAMD: Hamilton Depression Rating Scale.
Physiosom: physiosomatic, GIS: gastro-intestinal
Physio-affective core: first factor score extracted from pure and physiosom HAMD/HAMA and pure FF scores

### Construction of pathway-phenotypes

To construct pathway phenotypes, we employed factor analysis to examine whether latent vectors could be extracted from the SpO2 and body temperature data and the clinical rating scale scores. The results are shown in **Table 3**. The first FA was performed on SpO2, body temperature, TO2 index, and the 5 clinical scale subdomains. This data set showed a sufficient factorability of the correlation matrix and the first factor explained 64.19% of the variance and all factor loadings were > 0.66 with an adequate Cronbach alpha value. This factor, therefore, was dubbed the “TO2- physio-affective” or “TO2PA” pathway phenotype”. We could also extract a single latent vector from the SpO2, body temperature, TO2 index, chronic fatigue, GIS, sleep and autonomic symptoms with adequate KMO, Cronbach alpha, and explained variance data.

**Table 3:** Results of factor analysis (FA) conducted on body temperature, oxygen saturation (SpO2) and clinical rating scales

| **Features** | **FA#1** | **Features** | **FA#2** |
| --- | --- | --- | --- |
| TO2 index | **0.898** | TO2 index | **0.937** |
| SpO2 | **-0.821** | SpO2 | **-0.819** |
| Body temperature | **0.721** | Body temperature | **0.766** |
| Pure HAMD | **0.703** | Chronic fatigue | **0.750** |
| Physiosom HAMD | **0.809** | GIS | **0.674** |
| Pure HAMA | **0.662** | Sleep | **0.693** |
| Physiosom HAMA | **0.882** | Autonomic | **0.854** |
| Pure FF | **0.877** |  |  |
| KMO | 0.772 | KMO | 0.712 |
| % Variance | 64.19% | % Variance | 62.29% |
| Cronbach alpha | 0.784 | Cronbach alpha | 0.704 |

KMO: Keiser-Meier-Olkin test, SpO<sub>2</sub>: Oxygen saturation, FF: Fibro-fatigue scale, HAMD: Hamilton Depression Rating Scale, HAMA: Hamilton Anxiety Rating Scale. TO2 index: computed as $z \text{ body temperature} - z \text{ SpO}_2$ .

### Prediction of the clinical rating scales

We performed different multiple regression analyses using the subdomain scores as dependent variables and SpO2, body temperature, vaccination status (entered as dummy variables), age, sex, TUD, and education as explanatory variables (**Table 4**). Regression #1 shows that 38.9% of the variance in pure HAMD scores could be explained by SpO2, education, age (inversely) and body temperature (positively associated). Regression #2 shows that a large portion of the variance (42.7%) in Physiosom HAMD could be explained by SpO2 (inversely) and body temperature (positively) and being vaccinated with AstraZeneca or Pfizer. We found that (regression #3) 33.9% of the variance in pure HAMA was explained by a model involving SpO2 (negatively), female sex, and vaccination with AstraZeneca. The physiosom HAMA (regression #4) was best predicted by SpO2, body temperature, female sex and vaccination with AstraZeneca or Pfizer. Regression #5 shows that 54.9% of the variance in pure FF scores could be explained by SpO2 (inversely) and peak body temperature (positively). Regression #6 showed that 60.7% of the variance in the physio-affective phenome score was explained by SpO2 (inversely), peak body temperature, female sex and vaccination with AstraZeneca or Pfizer. **Figures 1 and 2** show the partial regression of the physio-affective phenome score on SpO2 and body temperature, respectively. **Figure 3** shows the partial regression of the physio-affective phenome on the TO2 index. Also, in the restricted study sample of patients with Long COVID we found that SpO2 levels were significantly correlated with Pure HAMD (r=0.258, p=0.005, n=120), Physiosom HAMD (r=0420, p<0.001), Pure HAMA (r=0.334, p<0.001), Physiosom HAMA (r=0.559, p<0.001), and Pure FF (r=0.463, p<0.001) scores. These effects remained significant using an FDR of p=0.01. After FDR p correction, no significant correlations were observed between body temperature and the clinical scale scores in the patient sample. In the restricted study sample of COVID patients, we found a significant association between the physio-affective phenome score and the TO2 index (r=0.519, p<0.001, n=118). **Figure 4** shows the partial regression of the physio-affective phenome on the TO2 index in the restricted study sample of COVID-19 patients only.

**Figure 1.**
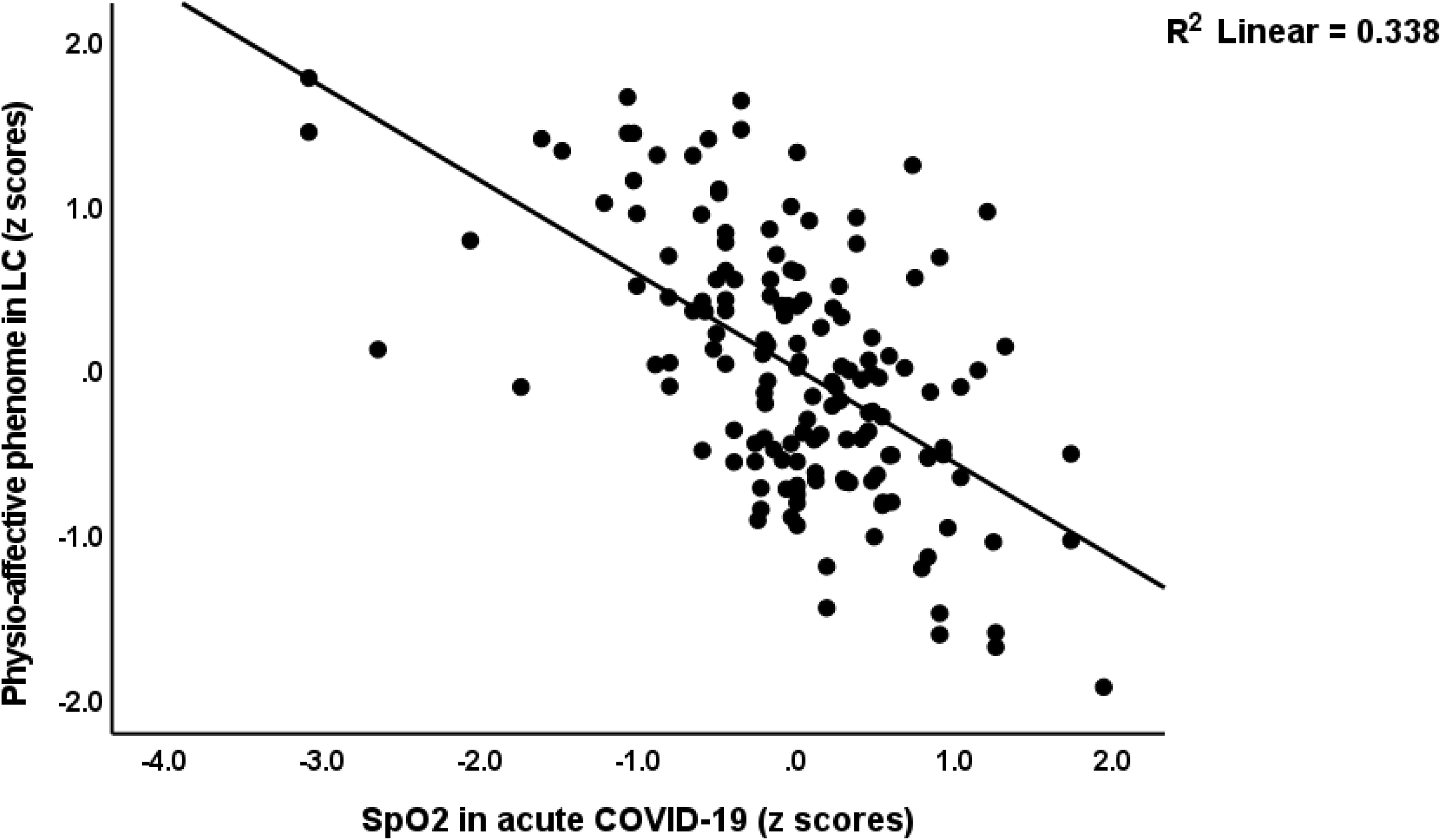
Partial regression of the physio-affecti.ve phenome score in controls and patients with Long COVID (LC) on oxygen saturation levels

**Figure 2.**
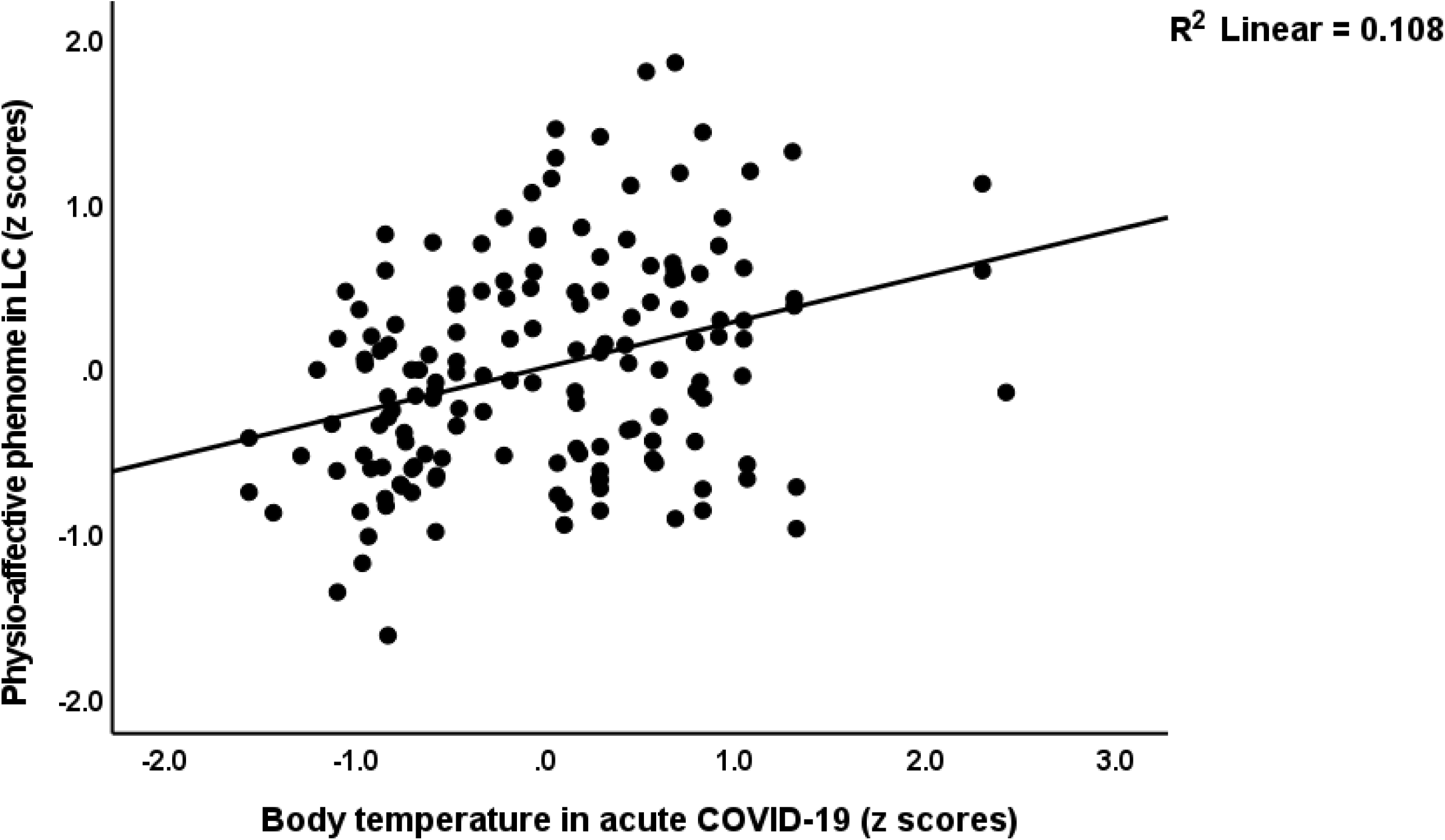
Partial regression of the physio-affecti.ve phenome score in controls and patients with Long COVID (LC) on peak body temperature

**Figure 3.**
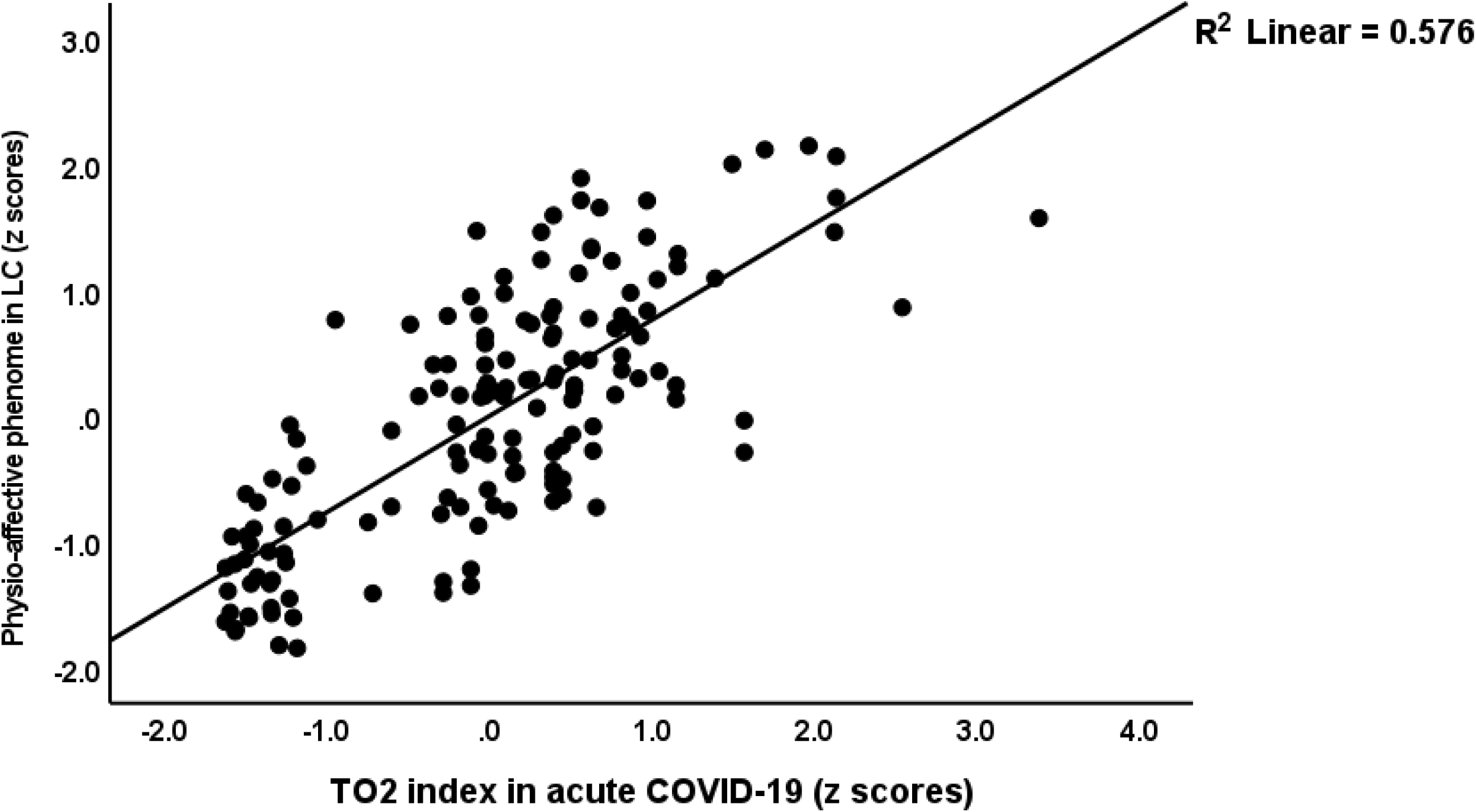
Partial regression of the physio-affecti.ve phenome score in controls and patients with Long COVID (LC) on the T02 index, which combines higher body temperature and lower oxygen saturation

**Figure 4.**
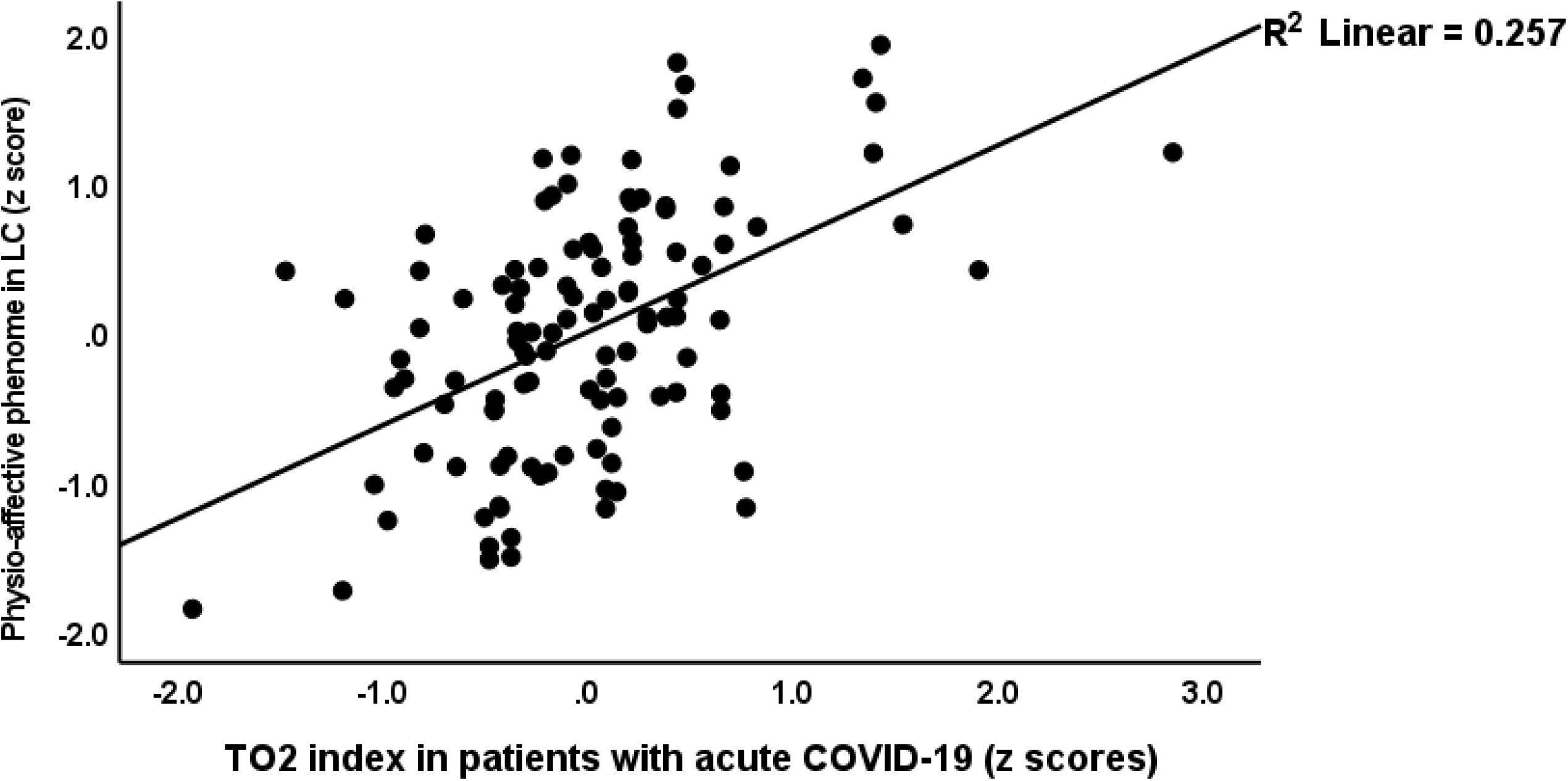
Partial regression of the physio-affecti.ve phenome score in patients with Long COVID (LC) on the T02 index during acute COVID-19, which combines higher body temperature and lower oxygen saturation

**Table 4:** Results of multiple regression analyses with psychiatric rating scales and subdomain scores as dependent variables.

| Dependent Variables | Explanatory Variables | Coefficients of input variables |  |  | Model statistics |  |  |  |
| --- | --- | --- | --- | --- | --- | --- | --- | --- |
| | | $\beta$ | t | p | R<sup>2</sup> | F | df | p |
| **#1. Pure HAMD** | **Model** |  |  |  | 0.389 | 23.67 | 4/149 | <0.001 |
|  | Body temperature | 0.387 | 4.86 | <0.001 |  |  |  |  |
|  | SpO2 | -0.268 | -3.36 | <0.001 |  |  |  |  |
|  | Education | -0.146 | -2.25 | 0.026 |  |  |  |  |
|  | Age | -0.135 | -2.10 | 0.038 |  |  |  |  |
| **#2. Physiosom HAMD** | **Model** |  |  |  | 0.427 | 37.20 | 3/150 | <0.001 |
|  | spO2 | -0.468 | -6.02 | <0.001 |  |  |  |  |
|  | Body temperature | 0.220 | 2.85 | 0.005 |  |  |  |  |
|  | AstraZeneca or Pfizer | 0.155 | 2.47 | 0.015 |  |  |  |  |
| **#3. Pure HAMA** | **Model** |  |  |  | 0.339 | 25.68 | 3/150 | <0.001 |
|  | spO2 | -0.511 | -7.58 | <0.001 |  |  |  |  |
|  | Female sex | 0.217 | 3.26 | 0.001 |  |  |  |  |
|  | AstraZeneca | 0.134 | 1.98 | 0.049 |  |  |  |  |
| **#4. Physiosom HAMA** | **Model** |  |  |  | 0.566 | 48.51 | 4/149 | <0.001 |
|  | spO2 | -0.565 | -8.31 | <0.001 |  |  |  |  |
|  | Body temperature | 0.228 | 3.38 | <0.001 |  |  |  |  |
|  | AstraZeneca or Pfizer | 0.134 | 2.45 | 0.015 |  |  |  |  |
|  | Female Sex | 0.120 | 2.22 | 0.028 |  |  |  |  |
| **#5. Pure FF** | **Model** |  |  |  | 0.549 | 91.91 | 2/151 | <0.001 |
|  | SpO2 | -0.515 | -7.60 | <0.001 |  |  |  |  |
|  | Body temperature | 0.309 | 4.57 | <0.001 |  |  |  |  |
| **#6. Physio-affective phenome score** | **Model** |  |  |  | 0.607 | 57.64 | 4/149 | <0.001 |
|  | SpO2 | -0.565 | -8.72 | <0.001 |  |  |  |  |
|  | Body temperature | 0.273 | 4.25 | <0.001 |  |  |  |  |
|  | Female sex | 0.115 | 2.23 | 0.027 |  |  |  |  |
|  | AstraZeneca or Pfizer | 0.112 | 2.15 | 0.033 |  |  |  |  |

SPO<sub>2</sub>: Oxygen saturation, FF: Fibro-Fatigue scale, HAMA: Hamilton Anxiety Rating Scale, HAMD: Hamilton Depression Rating Scale; Physio-affective phenome score: first factor score extracted from pure and physiosom HAMD/HAMA and pure FF scores

### Results of canonical correlations

To delineate the associations between SpO2 and body temperature and the different symptom profiles of Long COVID we used canonical correlation analysis with the Long COVID symptom profiles as dependent variables. Table 5 shows that a canonical component extracted from SpO2 and body temperature (explaining 76.6% of the variance) was strongly correlated (explaining 31.0% of the variance) with a factor extracted from HAMD symptoms (explaining 55.1% of the variance), namely depressed mood, insomnia early and middle, GIS and genital symptoms, and hypochondriasis. The same biomarkers explained 31.9% of the variance in a factor extracted from 9 FF symptoms, namely muscle pain and tension, fatigue, irritability, sleep disorders, autonomic and GIS symptoms, headache and a flu-like malaise. Baseline SpO2 and body temperature also explained 34.3% of the variance in a factor extracted from 8 HAMA symptoms, namely anxious mood, tension, insomnia, depressed mood, and sensory, respiratory, genitourinary and autonomic symptoms.

**Table 5.** Results of canonical correlation analyses examining the effects of body temperature and oxygen saturation (SpO2) on the mental and physiological symptoms of Long COVID

|  | HAMD |  | FF |  | HAMA |  |
| --- | --- | --- | --- | --- | --- | --- |
| Feature sets | Variables | C Loadings | variables | C Loadings | variable | C Loadings |
| **Set 1**<br>**Clinical** | Depressed mood | 0.672 | Muscle pain | 0.882 | Anxious mood | 0.583 |
|  | Insomnia early | 0.674 | Muscle tension | 0.744 | Tension | 0.759 |
|  | Insomnia middle | 0.818 | Fatigue | 0.835 | Insomnia | 0.684 |
|  | Somatic GIS | 0.681 | Irritability | 0.653 | Depressed mood | 0.635 |
|  | Genital symptoms | 0.858 | Sleep | 0.652 | Sensory | 0.627 |
|  | Hypochondriasis | 0.729 | Autonomic | 0.802 | Respiratory | 0.878 |
|  |  |  | GIS | 0.577 | Genitourinary | 0.723 |
|  |  |  | Headache | 0.747 | Autonomic | 0.858 |
|  |  |  | Malaise | 0.613 |  |  |
| **Set 2**<br>**Biomarkers** | Body temperature | 0.7675 | Body temperature | 0.787 | Body temperature | 0.749 |
|  | SpO2 | -0.971 | SpO2 | -0.963 | SpO2 | -0.977 |
| **Statistics** | F (df) | 14.07 (12/292) |  | 11.06 (18/286) |  | 15.59 (16/288) |
|  | p | <0.001 |  | <0.001 |  | <0.001 |
|  | Correlation | 0.750 |  | 0.771 |  | 0.807 |
|  | Set 1 by set 2 | 0.310 |  | 0.319 |  | 0.343 |
|  | Set 1 by self | 0.551 |  | 0.537 |  | 0.526 |
|  | Set 2 by self | 0.766 |  | 0.773 |  | 0.758 |

C Loadings: Canonical Loadings, GIS: gastro-intestinal symptoms, HAMD: Hamilton Depression Rating Scale; FF: Fibro-fatigue scale; HAMA: Hamilton Anxiety Rating Scale

## Discussion

### Clinical aspects of Long COVID

The first major finding of the current study is that increased body temperature and especially decreased levels of SpO2 in acute COVID-19 predict the onset of mental symptoms, chronic fatigue and physiosomatic (previously named psychosomatic) symptoms that characterize Long COVID. Moreover, based on these two baseline markers of acute COVID-19, we were able to construct a new endophenotype cluster of Long COVID patients who show very low SpO2, high body temperature, and increased levels of depressive, anxiety and physiosomatic symptoms, including autonomic and GIS, sleep disorders, fatigue, and cognitive impairments. The estimated number of patients in this new TO2PA (TO2-physio-affective) endophenotype class was around 26.7% of the Long COVID patients. We should stress that the current study did not aim to estimate the prevalence of Long COVID mental symptoms but rather to examine whether baseline biomarkers of infection and immune activation predict mental symptoms and, using the precision nomothetic approach (Maes 2022) to define new endophenotype classes and pathway phenotypes to examine the pathophysiology of Long COVID.

The current results extend those of previous reports, which ubiquitously reported mental and physiosomatic symptoms in Long COVID patients (Huang, Huang et al. 2021, Taquet, Geddes et al. 2021, Titze-de-Almeida, da Cunha et al. 2022). Moreover, recent meta-analyses revealed that the top symptoms of Long COVID were in descending order of importance: fatigue, brain fog, memory disturbances, attention problems, myalgia, anosmia, dysgeusia, and headache (Premraj, Kannapadi et al. 2022). Similar findings were reported in another meta-analysis (Badenoch, Rengasamy et al. 2022) showing that the top most prevalent symptoms were in descending order of importance: sleep disturbances, fatigue, objective cognitive deficits, anxiety and post-traumatic stress. Moreover, these meta-analyses showed that the prevalence of mental symptoms including depression tends to increase over the time from mid to long-term follow up (Premraj, Kannapadi et al. 2022).

Previously, we observed that the acute infectious phase was characterized by intertwined increases in key depression, anxiety and physiosomatic symptoms as assessed with the HAMD, HAMA and FF scales (Al-Jassas, Al-Hakeim et al. 2022). As such, both acute COVID-19 and Long COVID are accompanied by significant intertwined increases in mental and chronic fatigue symptoms. Furthermore, both in the acute infectious phase and Long COVID one single latent trait could be extracted from these mental and physiosomatic symptoms indicating that these symptoms are manifestations of a common core, namely the “physio-affective phenome” of COVID-19 and Long COVID. This indicates that shared pathways may underpin the physio-affective phenome of the acute as well as chronic phases of the illness. Previously, we observed intertwined associations between increased levels of affective and physiosomatic symptoms not only in acute COVID-19 but also in, for example, schizophrenia, rheumatoid arthritis, and major depression (Kanchanatawan, Sriswasdi et al. 2019, Almulla, Al-Hakeim et al. 2020, Maes, Andrés-Rodríguez et al. 2021, Smesam, Qazmooz et al. 2022). Since our previous study (Al-Jassas, Al-Hakeim et al. 2022) and the current study were performed using different study samples, we were unable to examine whether patients with acute physio-affective symptoms present the same symptoms in Long COVID. Nevertheless, since we excluded in both studies patients with primary major depression, anxiety disorders and chronic fatigue syndrome, our findings indicate that SARS-CoV- 2 infected patients develop de novo mental symptoms and chronic fatigue during both the acute and the chronic phase of the illness.

### Biomarkers of acute COVID-19 and Long COVID

The second major finding of this study is that a large part of the severity of the physio-affective core (60.7%) during Long COVID was significantly predicted by SpO2 and body temperature values during the acute phase of the disease. In the latter, we observed that the physio-affective core was strongly associated with a replicable latent vector extracted from SpO2, CCTAs (including crazy patterns, consolidation, ground glass opacities), increased levels of pro-inflammatory and anti-inflammatory cytokines, and SARS-Cov2 infection (Al-Jassas et al., 2022). These findings indicate that during the acute phase of illness, lowered SpO2 is a manifestation of the infection-immune-inflammatory core which is accompanied by CCTAs. As reviewed in the Introduction, the degree of increased body temperature in the acute phase reflects the severity of inflammation. Moreover, for every 0.5 °C increase in body temperature there is an increase in mortality rate reaching 42.0% in people with a body temperature > 40.0 °C (Tharakan et al., 2020). As such, increased body temperature not only predicts increased mortality rates but also increased severity of the physio-affective phenome.

It should be stressed that during the initial phase of COVID-19 infection, a sickness behavioral complex (SBC) is present, which includes physiosomatic symptoms such as muscle pain and tension, loss of appetite, fatigue, headache and probably also dysgeusia and anosmia (Maes, Tedesco Junior et al. 2022). This SBC protects against severe and critical COVID-19 disease and is partly mediated by NLRP3 (nucleotide-binding domain, leucine-rich repeat and pyrin domain-containing protein 3 inflammasome) gene variants (Maes, Tedesco Junior et al. 2022). Nevertheless, the SBC is a beneficial short-lasting response confined to the acute phase of inflammation and should be discriminated from the affective and chronic fatigue symptoms which accompany the chronic inflammatory phase (Maes, Berk et al. 2012, Morris, Anderson et al. 2013). Our findings that lowered levels of SpO2 and increased body temperature (and consequently also CCTAs and inflammation) are associated with Long COVID physio-affective symptoms may be explained by several factors. First, both increased body temperature and lowered SpO2 during the acute phase indicate more severe inflammatory responses (Tharakan, Nomoto et al. 2020, Al-Jassas, Al-Hakeim et al. 2022), which could further develop into chronic inflammatory responses (Maes, Berk et al. 2012). Signs of activated immune-inflammatory pathways were observed in Long COVID including increased levels of interleukin (IL)-2, IL-1β, IL-6, IL-17A, IL-12p70, interferon (IFN)-γ, tumor necrosis factor (TNF)-α and macrophage inflammatory protein1β, and increased levels of acute phase reactants such as C-reactive protein and ferritin (Breton, Mendoza et al. 2020, Mazza, De Lorenzo et al. 2020, Santis, Pérez-Camacho et al. 2020, García-Abellán, Padilla et al. 2021, Ong, Fong et al. 2021, Sonnweber, Sahanic et al. 2021, Ceban, Ling et al. 2022). Activation of immune-inflammatory pathways may explain the onset of affective and physiosomatic symptoms as well as chronic fatigue syndrome (Maes, Berk et al. 2012, Morris, Anderson et al. 2013).

Second, lowered SpO2 itself may cause fatigue and depressive symptoms (Pan, Zhao et al. 2015, Zhao, Yang et al. 2017) and is implicated in cognitive impairments (Wang, Cui et al. 2022), autonomic symptoms (Chen, Chen et al. 2006) and insomnia (Johansson, Svensson et al. 2015). Hypoxia-inducible factors (HIFs) are key regulators of oxygen homeostasis (Yoon, Pastore et al. 2006) which are induced in response to hypoxia thereby promoting angiogenesis (Carmeliet, Dor et al. 1998) and anaerobic metabolism (Carmeliet, Dor et al. 1998, Vaupel 2004), while lowering mitochondrial oxygen via activating pyruvate kinase I enzyme and inhibiting the citric acid cycle (Ziello, Jovin et al. 2007, Morris, Maes et al. 2019). Importantly, HIF1A is part of the immune protein-protein interaction network of affective disorders (Maes, Rachayon et al. 2022) and inflammatory responses in general (Cramer, Yamanishi et al. 2003, Oda, Hirota et al. 2006, Imtiyaz and Simon 2010). Hence, hypoxia and inflammation in acute COVID-19 may be accompanied by overexpression of HIFs which may further fuel the immune-inflammatory disorders leading to Long COVID. Moreover, hypoxia may cause increases in reactive oxygen and nitrogen species (Solaini, Baracca et al. 2010), leading to oxidative damage, which is implicated in the pathophysiology of depression, fatigue and anxiety (Maes, Kubera et al. 2011, Morris and Maes 2014). Furthermore, different areas of the brain, mainly the structures that take part in affective disorders, namely the amygdala, hippocampus, anterior cingulate cortex, and prefrontal cortex (Aryutova and Stoyanov 2021) were found to be influenced by hypoxia (Shankaranarayana Rao, Raju et al. 1999, Alchanatis, Zias et al. 2005).

Third, decreased SpO2 in acute COVID-19 is attributed to lung inflammation, bronchitis, pneumonia and lung fibrosis as indicated by the presence of CCTAs (Sadhukhan, Ugurlu et al. 2020, Solomon, Heyman et al. 2021, Al-Jassas, Al-Hakeim et al. 2022). Up to fifty percent of the post-COVID-19 patients may show some signs of lung fibrosis (Nabahati, Ebrahimpour et al. 2021) and 2-6% of Long COVID patients who experienced moderate COVID-19 illness develop lung fibrosis (Bazdyrev, Rusina et al. 2021). In addition, a significant cohort of recovered patients show more persistent lung inflammation which may cause physiological and functional changes (Myall, Mukherjee et al. 2021) and even CCTAs were reported in some of Long COVID patients (Solomon, Heyman et al. 2021, Vijayakumar, Tonkin et al. 2021). All in all, increased lung inflammation and fibrosis in the post-infectious phase may further contribute to lowered SpO2 and immune-inflammatory responses and thus the physio-affective phenome of Long COVID. A fourth possibility is that some COVID vaccines contribute to the physio-somatic phenome of Long COVID. In this regard we observed that AstraZeneca and Pfizer vaccinations aggravated the physiosomatic phenome, whereas Sinopharm had no such effect.

## Limitations

Some limitations and strengths should be considered while interpreting the current results. First, the paper would have been more interesting if we had measured HIFs and the tryptophan catabolite (TRYCAT) pathway in the acute and chronic phase of the disease. Indeed, a recent meta-analysis showed that neurotoxic TRYCATs are significantly increased in acute COVID-19, while TRYCATs are known to be associated with the onset of affective, physiosomatic and cognitive symptoms (Maes, Leonard et al. 2011, Kanchanatawan, Sirivichayakul et al. 2018, Almulla and Maes 2022, Almulla, Supasitthumrong et al. 2022). Second, although we conducted a case-control study, we also measured body temperature and SpO2 in the acute phase of illness using a retrospective cohort study design which allows to examine causal associations.

## Conclusions

In people with Long COVID, low SpO2 and higher peak body temperature during the acute phase predict the affective and physiosomatic symptoms, chronic fatigue, sleep disturbances, cognitive impairments, and GIS and autonomic symptoms of Long COVID. As such, lowered SpO2 and higher body temperature and the associated CCTAs and immune-inflammatory responses during the acute phase are new drug targets to prevent the Long COVID-associated physio-affective phenome.

## Data Availability

All data produced in the present work are contained in the manuscript

## Acknowledgements

The authors thank the staff of Al-Sader Medical City of Najaf, Al-Hakeem General Hospital, Al-Zahraa Teaching Hospital for Maternity and Pediatrics, Imam Sajjad Hospital, Hassan Halos Al-Hatmy Hospital for Transmitted Diseases, Middle Euphrates Center Cancer, Al-Najaf Center for Cardiac Surgery and Trans Catheter Therapy for their efforts in the collection of data.

## Ethical approval and consent to participate

All the controls and patients or their parents/legal guardians provided written signed consent. The approval of the study was obtained from the institutional ethics board of the University of Kufa (617/2020). The study was conducted according to Iraqi and foreign ethics and privacy laws in accordance with the guidelines of the World Medical Association Declaration of Helsinki, The Belmont Report, CIOMS Guideline, and International Conference on Harmonization of Good Clinical Practice; our IRB adheres to the International Guideline for Human Research Safety (ICH-GCP).

## Declaration of interest

The authors declare no financial conflict of interests.

## Funding

There is no specific funding for this study

## Author’s contributions

The preparation of the manuscript was made with participation of all authors and they approved the final version.

## Notes

### Competing Interest Statement

The authors have declared no competing interest.

